## Supplemental Material for "Quantifying the spatiotemporal dynamics of the first two epidemic waves of SARS-CoV-2 infections in the United States"

**Table of Figures:**

**Figure 1: United States population and estimated cumulative SARS-CoV-2 infections per 100,000 distributed across the hexagonal grid.**

**Figure 2: Estimated infections per 100,000 of SARS-CoV-2 in the United States, March 2020–December 2021.**

**Figure 3: Progression of the wave boundary measured as the distance to the nearest point on the boundary at t+1 (in km/day).**

**Figure S1 – Histogram of values for the risk surfaces and empirical cumulative density function of the risk surface values.**

**Figure S2 – Sensitivity Analysis of recruitment rate to the threshold values.**

**Figure S3 – Sensitivity analysis maps to the threshold value, no threshold**

**Figure S4 – Sensitivity analysis maps to the threshold value, threshold is 85**

**Table S5 – Daily estimates of wave length, areal growth, and speed for each wave**

**Prior choice for the modified Besag-York-Mollié model, BYM2**

We fitted the BYM2 model using R-INLA(1). The INLA methodology represents an approximation to full Markov chain Monte Carlo sampling algorithms, which can drastically reduce computational times, particularly for large analysis datasets. The original BYM model combines the Besag (7) version of an intrinsic conditional autoregressive (ICAR) model with an independent error term to balance spatial smoothness with non-spatial variability. BYM models use an adjacency matrix that defines regions as 'neighbors' based on whether they have touching boundaries. BYM2 is a reparametrized version of this model that leads to improvements when assigning prior distributions for model parameters and improved interpretations of those parameters (6). The BYM2 implementation in the R-INLA package makes use of penalized-complexity (PC) priors, which implies a risk decomposition as suggested by MacNab, 2011. From our model defined in (1) and following the reparameterization in Riebler et al., 2016 we have for $\theta(A_i)$,

$$\theta(A_i) = \frac{1}{\tau_b}(\sqrt{1-\phi}\,v(A_i) + \sqrt{\phi}\,u(A_i) \quad (1)$$

where $\tau_b$ is a precision parameter and $\phi$ is the mixing parameter. Here, the $v(A_i)$ are *iid* with a normal distribution and variance equal to one, and the $u(A_i)$ parameters are assigned the Besag intrinsic conditional autoregressive (ICAR) model with variance equal to 1.

This reparameterization of the BYM model can be seen as a mixture between pure overdispersion (when $\phi = 0$), and a completely spatially structured risk (when $\phi = 1$ the Besag model). We used a conservative PC-prior for $\phi$, specifically, P($\phi > 1/2 = 2/3$) that assumes the unstructured random effect accounts for more variability than the spatially structured effect. For the precision parameter, $\tau_b$, we also used a PC-prior with parameterization P($\tau_b > 0.2 = 0.01$).

**Sensitivity Analysis on infections per 100,000 surface threshold**

In the section **Definition of wave and speed of expansion of waves**, we define the threshold for a high rate of infection per 100,000 surfaces in the speed of invasion calculation as any value above the 75$^{th}$ percentile (**Figure S1**, Panel **B**) of the surface values distribution, which corresponds to a value of 190 or more infections per 100,000. To test whether this threshold changes the assessment of per 100,000 infection surfaces and the wave-like patterns visualization, we conducted the following sensitivity analysis. First, we inspected the histogram of values of the infection rate surface for all the dates over all the hexes, **Figure S1A.** We found that most of the mass of the distribution, disregarding the values close to 0, is around 200 infections per 100,000, with a steep decrease to values greater than 233 infections per 100,000. To further check if the value of 190 infections per 100,000 is a reasonable choice for the threshold, we inspected the empirical cumulative density function (ECDF) of the denoised and spatially smoothed random effects (**Figure S1B**). Setting the threshold to a value equal to or greater than 190 infections per 100,000 which we think is the best choice. At much lower threshold values, deviation of values close to 0 will put more hexagons as being part of a wave. If the value of infections per 100,000 is greater than 190, we conclude from the shape of the ECDF function that it will be hard to classify any region as having a high rate of infection. Of course, the value can be set to bigger or smaller values than 190 infections per 100,000, but the wave-like pattern does not disappear. **Figure S2** shows the speed of invasion for both waves, calculated with different thresholds, 127 or more infections per 100,000 for panel **A** and 233 infections per 100,000 for panel **B**. Those values are the median and the 90$^{th}$ percentile of the distribution.

To check whether the wave-like pattern is sensitive to the choice of threshold, we recreated **Figure 2** for the same eight dates using a continuous scale (**Figure S3)** or by setting the threshold to the median of the infections per 100,000 distribution (127 infections per 100,000)(**Figure S4**).

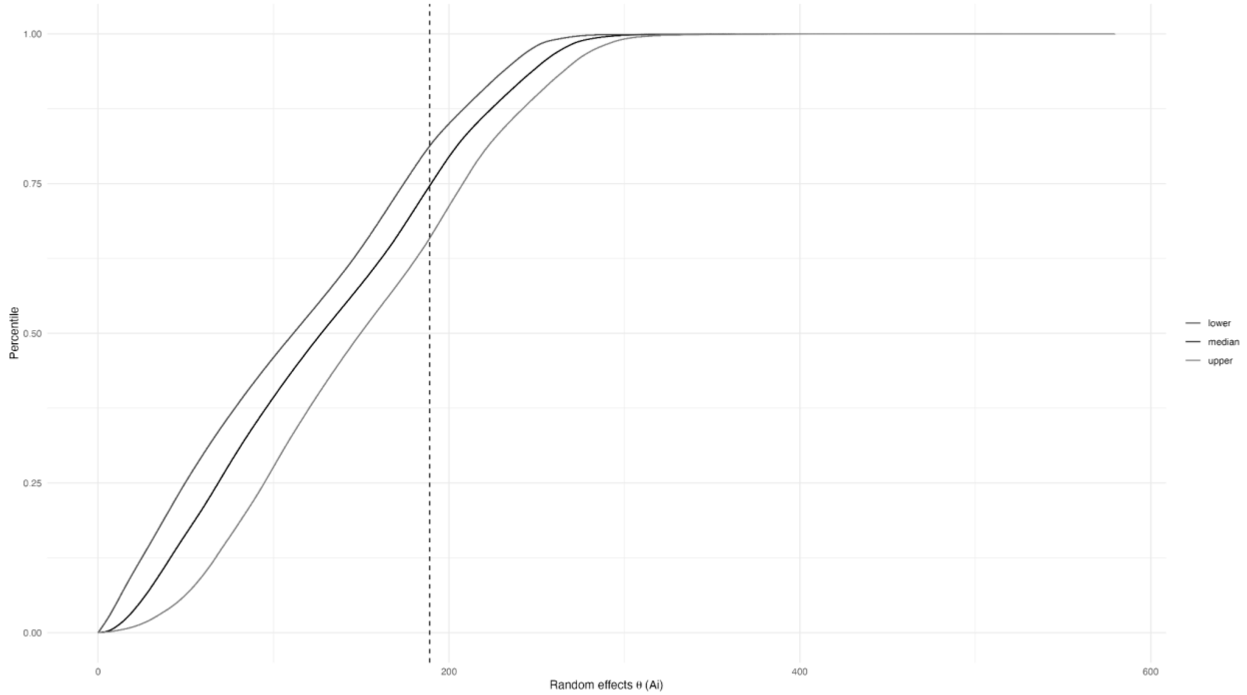


**Figure S1 – Empirical cumulative density function (ECDF) of the risk surface values.** The ECDF shows that a threshold of 190 infections per 100,000 is indicated by the vertical dashed line at which the ECDF crosses the 75th percentile. The ECDF for the lower and upper bounds is shown in grey.

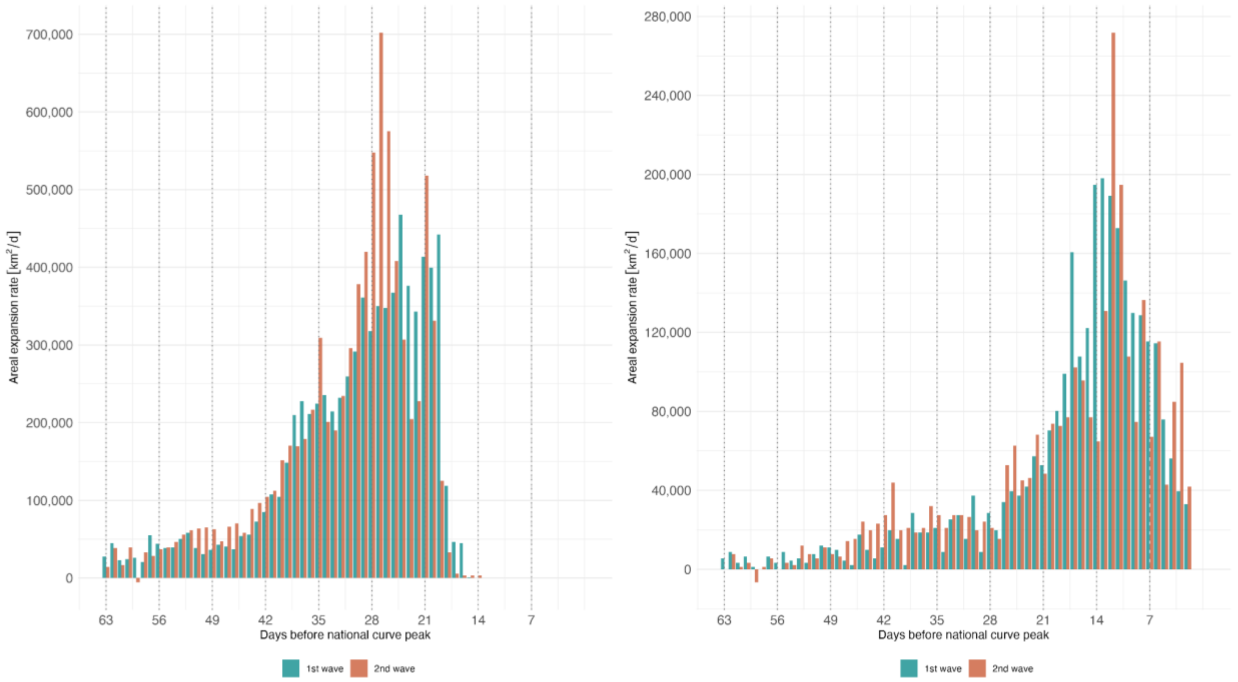


**Figure S2 – Sensitivity analysis of areal wave expansion to the threshold values.** Areal wave expansion (km2/day) for different thresholds of infection per 100,00 in the progression calculation of the surfaces. Panel **A** is built with a threshold of 127 or more infections per 100,000; Panel **B** is built with a threshold of 233 infections per 100,000. As in **Figure 3C**, we observe a maximal speed and a steep decrease after the peak, and the second wave had a higher invasion speed and encompassed a larger area at peak than the first wave.

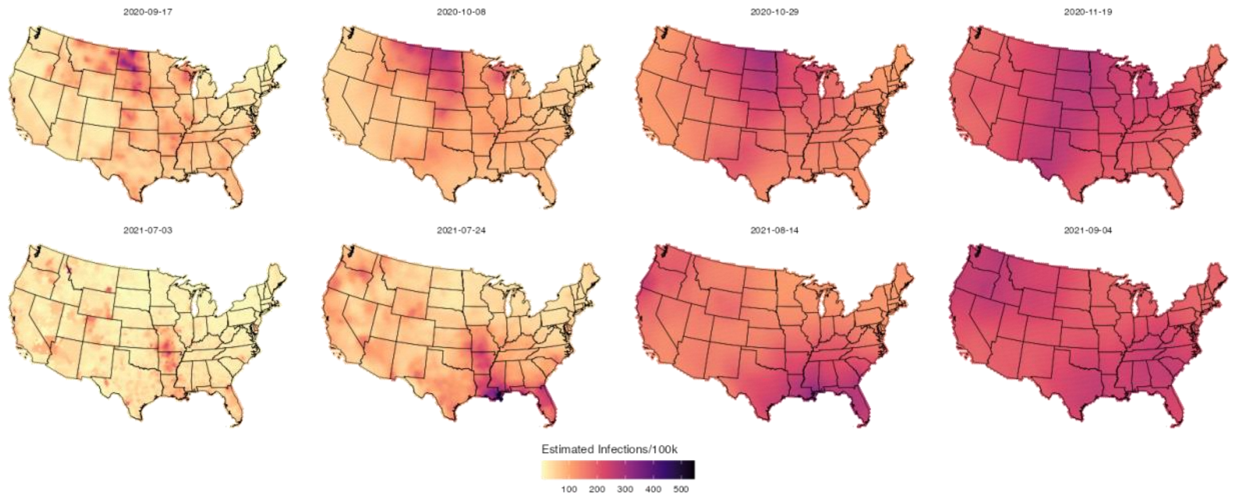


**Figure S3** – Infection per 100,000 persons surface on a continuous scale of values. As expected, the wave-like pattern holds independently of the scale to be displayed, and as being an output of spatial smooth model, the continuous scale gives a less defined border to the risk surface expansion. All maps were generated using United States, state, and county borderlines maps in public domain from the Census Bureau which were downloaded through the R package Tigris (30). The shapefile generated for this analysis with the population estimates and cumulative infections estimates can be found at: https://github.com/covidestim/waves/tree/waves-manuscript/Data/data-products.

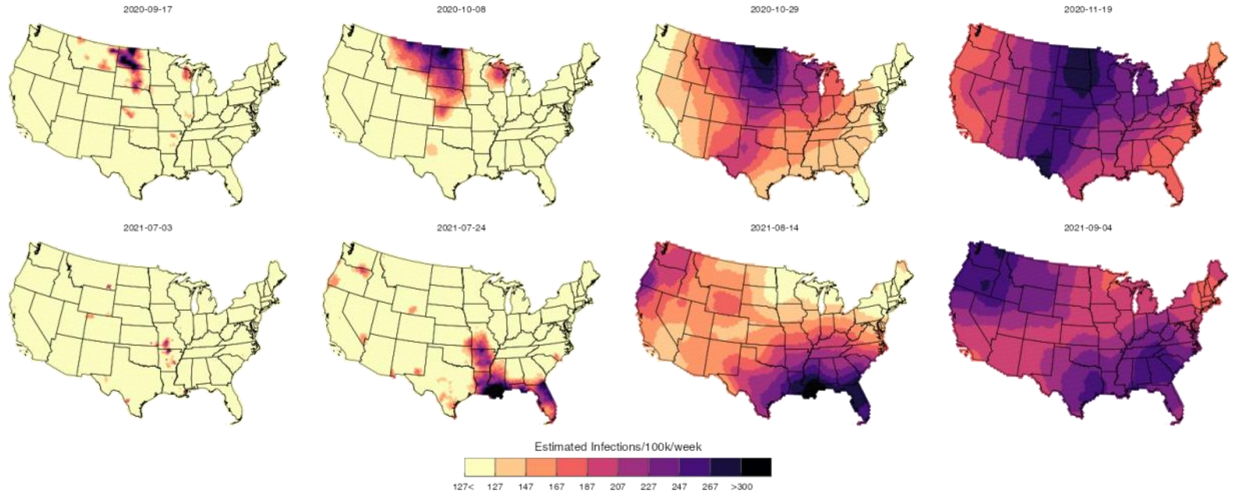


**Figure S4** - Infection per 100,000 with a threshold equal to the mean of the risk values distribution (127 infections per 100,000). With a lower threshold showing on the map, the spread process seems to happen faster. All maps were generated using United States, state, and county borderlines maps in the public domain from the Census Bureau, which were downloaded through the R package Tigris (30). The shapefile generated for this analysis with the population estimates and cumulative infections estimates can be found at: https://github.com/covidestim/waves/tree/waves-manuscript/Data/data-products.

**S1_Table – Daily estimates of wave length, areal growth, and speed for each wave**

| | Wave 1 | | | | | Wave 2 | | | | |
|---|---|---|---|---|---|---|---|---|---|---|
| Days before peak | Wave edge length (km) | Areal wave expansion (km2/day) | Median speed (km/day) | Mean speed (km/day) | Length of wave edge with speed greater than or equal to overall median max speed | Wave edge length (km) | Areal wave expansion (km2/day) | Median speed (km/day) | Mean speed (km/day) | Length of wave edge with speed greater than or equal to overall median max speed |
| 63 | 3251.1 | 12100.0 | 0.0 | 2.3 | 535.0 | 1605.0 | 1100.0 | 0.0 | 2.6 | 164.6 |
| 62 | 3436.3 | 6600.0 | 0.0 | 1.8 | 391.0 | 1769.6 | 16500.0 | 0.0 | 1.4 | 205.8 |
| 61 | 3395.1 | 12100.0 | 0.0 | 3.4 | 637.9 | 2860.1 | 1100.0 | 0.0 | 0.7 | 123.5 |
| 60 | 3354.0 | 13200.0 | 0.0 | 2.3 | 535.0 | 2983.6 | 2200.0 | 0.0 | 33.6 | 370.4 |
| 59 | 3724.3 | 4400.0 | 0.0 | 1.0 | 246.9 | 3065.9 | -9900.0 | 0.0 | 21.9 | 946.5 |
| 58 | 3868.4 | 16500.0 | 0.0 | 3.5 | 864.2 | 2263.4 | 4400.0 | 0.0 | 0.6 | 102.9 |
| 57 | 3868.4 | 20900.0 | 0.0 | 3.9 | 905.4 | 2510.3 | 8800.0 | 0.0 | 1.0 | 185.2 |
| 56 | 4074.1 | 18700.0 | 0.0 | 4.3 | 967.1 | 2963.0 | 16500.0 | 0.0 | 1.3 | 288.1 |
| 55 | 3868.4 | 23100.0 | 0.0 | 3.8 | 864.2 | 3765.5 | 20900.0 | 0.0 | 2.5 | 720.2 |
| 54 | 4156.4 | 19800.0 | 0.0 | 5.8 | 967.1 | 4094.7 | 14300.0 | 0.0 | 1.8 | 555.6 |
| 53 | 4094.7 | 26400.0 | 0.0 | 4.3 | 1028.8 | 4341.6 | 23100.0 | 0.0 | 5.2 | 1028.8 |

| | | | | | | | | | | |
|---|---|---|---|---|---|---|---|---|---|---|
| 52 | 4485.7 | 20900.0 | 0.0 | 3.5 | 987.7 | 4444.5 | 12100.0 | 0.0 | 3.6 | 658.4 |
| 51 | 4568.0 | 18700.0 | 0.0 | 3.2 | 884.8 | 4609.1 | 22000.0 | 0.0 | 3.2 | 946.5 |
| 50 | 4691.4 | 20900.0 | 0.0 | 3.9 | 987.7 | 4938.3 | 28600.0 | 0.0 | 4.9 | 1152.3 |
| 49 | 4732.6 | 31900.0 | 0.0 | 4.8 | 1358.0 | 5041.2 | 17600.0 | 0.0 | 3.1 | 925.9 |
| 48 | 5103.0 | 28600.0 | 0.0 | 5.8 | 1460.9 | 5205.8 | 29700.0 | 0.0 | 6.0 | 1275.7 |
| 47 | 4917.8 | 16500.0 | 0.0 | 4.6 | 1111.1 | 5329.3 | 46200.0 | 0.0 | 13.9 | 2140.0 |
| 46 | 4670.9 | 12100.0 | 0.0 | 2.6 | 720.2 | 5123.5 | 47300.0 | 0.0 | 6.9 | 1522.7 |
| 45 | 4609.1 | 24200.0 | 0.0 | 5.1 | 1440.4 | 5473.3 | 44000.0 | 0.0 | 10.9 | 1995.9 |
| 44 | 4526.8 | 16500.0 | 0.0 | 4.7 | 1172.9 | 5082.4 | 37400.0 | 0.0 | 7.1 | 1666.7 |
| 43 | 4465.1 | 17600.0 | 0.0 | 3.3 | 967.1 | 5041.2 | 30800.0 | 0.0 | 5.6 | 1399.2 |
| 42 | 4423.9 | 18700.0 | 0.0 | 4.4 | 1275.7 | 5020.7 | 37400.0 | 0.0 | 7.1 | 1934.2 |
| 41 | 4341.6 | 25300.0 | 0.0 | 5.7 | 1440.4 | 5103.0 | 35200.0 | 0.0 | 18.5 | 2242.8 |
| 40 | 4259.3 | 15400.0 | 0.0 | 3.9 | 1090.6 | 4547.4 | 29700.0 | 0.0 | 8.4 | 1687.3 |
| 39 | 4259.3 | 16500.0 | 0.0 | 5.8 | 1522.7 | 4526.8 | 47300.0 | 0.0 | 13.7 | 2469.2 |
| 38 | 4115.3 | 36300.0 | 0.0 | 7.1 | 1749.0 | 4053.6 | 29700.0 | 0.0 | 11.9 | 1666.7 |
| 37 | 4238.7 | 35200.0 | 0.0 | 6.7 | 1543.2 | 3991.8 | 33000.0 | 0.0 | 7.9 | 1707.8 |
| 36 | 4423.9 | 44000.0 | 0.0 | 8.1 | 2140.0 | 4053.6 | 57200.0 | 0.0 | 12.9 | 2345.7 |
| 35 | 4547.4 | 47300.0 | 0.0 | 8.3 | 1975.3 | 4012.4 | 45100.0 | 0.0 | 9.7 | 1913.6 |
| 34 | 4794.3 | 35200.0 | 0.0 | 7.9 | 2037.1 | 4135.9 | 40700.0 | 0.0 | 8.4 | 1749.0 |
| 33 | 4588.5 | 66000.0 | 10.3 | 12.9 | 2839.5 | 4362.2 | 47300.0 | 0.0 | 11.6 | 2325.1 |

| | | | | | | | | | | |
|---|---|---|---|---|---|---|---|---|---|---|
| 32 | 4732.6 | 64900.0 | 20.6 | 12.5 | 2880.7 | 4423.9 | 69300.0 | 20.6 | 13.2 | 2716.1 |
| 31 | 4691.4 | 44000.0 | 0.0 | 8.0 | 2181.1 | 4670.9 | 78100.0 | 20.6 | 17.8 | 2880.7 |
| 30 | 4712.0 | 81400.0 | 20.6 | 16.2 | 3436.3 | 4568.0 | 82500.0 | 20.6 | 19.2 | 2963.0 |
| 29 | 4794.3 | 74800.0 | 20.6 | 13.4 | 3024.7 | 4691.4 | 107800.0 | 20.6 | 26.1 | 3498.0 |
| 28 | 4938.3 | 94600.0 | 20.6 | 18.7 | 3518.6 | 5144.1 | 128700.0 | 20.6 | 22.0 | 3868.4 |
| 27 | 5144.1 | 117700.0 | 20.6 | 21.7 | 3950.7 | 5432.2 | 114400.0 | 20.6 | 22.5 | 4012.4 |
| 26 | 5329.3 | 174900.0 | 20.6 | 42.4 | 4465.1 | 5185.3 | 139700.0 | 20.6 | 27.1 | 4362.2 |
| 25 | 5288.1 | 168300.0 | 20.6 | 29.6 | 4547.4 | 5247.0 | 174900.0 | 35.6 | 42.4 | 4609.1 |
| 24 | 6008.3 | 178200.0 | 20.6 | 31.6 | 5308.7 | 5103.0 | 169400.0 | 20.6 | 31.3 | 4177.0 |
| 23 | 5967.2 | 195800.0 | 35.6 | 30.4 | 5432.2 | 5699.7 | 172700.0 | 20.6 | 32.5 | 4814.9 |
| 22 | 6461.0 | 226600.0 | 35.6 | 34.8 | 5905.4 | 6193.5 | 229900.0 | 35.6 | 33.3 | 5720.3 |
| 21 | 6378.7 | 203500.0 | 35.6 | 34.4 | 6090.6 | 6769.7 | 190300.0 | 20.6 | 29.0 | 5823.1 |
| 20 | 6193.5 | 216700.0 | 35.6 | 32.5 | 5802.6 | 6378.7 | 177100.0 | 20.6 | 29.8 | 5617.4 |
| 19 | 6831.4 | 292600.0 | 35.6 | 40.5 | 6646.2 | 6214.1 | 204600.0 | 20.6 | 27.2 | 5411.6 |
| 18 | 7448.7 | 277200.0 | 35.6 | 50.5 | 7016.6 | 7592.7 | 217800.0 | 20.6 | 30.7 | 6769.7 |
| 17 | 6028.9 | 207900.0 | 35.6 | 37.3 | 5864.3 | 7777.9 | 283800.0 | 35.6 | 34.8 | 7078.3 |
| 16 | 5864.3 | 207900.0 | 35.6 | 38.3 | 5535.1 | 8518.6 | 297000.0 | 35.6 | 33.3 | 7736.7 |
| 15 | 5823.1 | 224400.0 | 35.6 | 44.0 | 5576.2 | 9424.0 | 332200.0 | 35.6 | 38.1 | 8765.6 |
| 14 | 5884.9 | 233200.0 | 35.6 | 44.0 | 5740.8 | 9259.4 | 336600.0 | 35.6 | 50.0 | 8889.0 |
| 13 | 5884.9 | 232100.0 | 35.6 | 43.6 | 5720.3 | 8703.8 | 345400.0 | 35.6 | 42.3 | 8251.2 |

| | | | | | | | | | | |
|---|---|---|---|---|---|---|---|---|---|---|
| 12 | 5946.6 | 246400.0 | 35.6 | 44.2 | 5843.7 | 8724.4 | 289300.0 | 20.6 | 36.4 | 7901.4 |
| 11 | 6111.2 | 260700.0 | 35.6 | 43.0 | 5946.6 | 8415.8 | 257400.0 | 20.6 | 29.9 | 7736.7 |
| 10 | 6522.7 | 225500.0 | 35.6 | 37.5 | 6378.7 | 8683.3 | 328900.0 | 35.6 | 37.6 | 8292.3 |
| 9 | 6502.2 | 234300.0 | 35.6 | 36.1 | 6234.7 | 9362.3 | 502700.0 | 54.4 | 74.6 | 9156.5 |
| 8 | 6893.1 | 239800.0 | 35.6 | 36.2 | 6646.2 | 8395.2 | 281600.0 | 35.6 | 50.4 | 7757.3 |
| 7 | 6934.3 | 233200.0 | 35.6 | 36.8 | 6769.7 | 6605.0 | 265100.0 | 35.6 | 71.4 | 6543.3 |
| 6 | 6522.7 | 122100.0 | 20.6 | 18.4 | 5061.8 | 5432.2 | 112200.0 | 20.6 | 23.8 | 4300.5 |
| 5 | 6502.2 | 187000.0 | 20.6 | 29.6 | 5864.3 | 4979.5 | 194700.0 | 35.6 | 45.4 | 4958.9 |
| 4 | 6317.0 | 112200.0 | 20.6 | 16.3 | 4465.1 | 4465.1 | 106700.0 | 20.6 | 24.9 | 3786.1 |
| 3 | 6193.5 | 108900.0 | 20.6 | 17.6 | 4279.9 | 4238.7 | 128700.0 | 35.6 | 32.4 | 3909.5 |
| 2 | 5926.0 | 101200.0 | 20.6 | 15.6 | 4177.0 | 4033.0 | 223300.0 | 54.4 | 60.1 | 4033.0 |
| 1 | 5823.1 | 71500.0 | 0.0 | 11.5 | 3333.4 | 3600.9 | 190300.0 | 35.6 | 42.5 | 3148.2 |
